# Designing isolation guidelines for COVID-19 patients utilizing rapid antigen tests: a simulation study using viral dynamics models

**DOI:** 10.1101/2022.01.24.22269769

**Authors:** Yong Dam Jeong, Keisuke Ejima, Kwang Su Kim, Woo Joohyeon, Shoya Iwanami, Yasuhisa Fujita, Il Hyo Jung, Kenji Shibuya, Shingo Iwami, Ana I. Bento, Marco Ajelli

## Abstract

Appropriate isolation guidelines for COVID-19 patients are warranted. Currently, isolating for fixed time is adapted in most countries. However, given the variability in viral dynamics between patients, some patients may no longer be infectious by the end of isolation (thus they are redundantly isolated), whereas others may still be infectious. Utilizing viral test results to determine ending isolation would minimize both the risk of ending isolation of infectious patients and the burden due to redundant isolation of noninfectious patients. In our previous study, we proposed a computational framework using SARS-CoV-2 viral dynamics models to compute the risk and the burden of different isolation guidelines with PCR tests. In this study, we extend the computational framework to design isolation guidelines for COVID-19 patients utilizing rapid antigen tests. Time interval of tests and number of consecutive negative tests to minimize the risk and the burden of isolation were explored. Furthermore, the approach was extended for asymptomatic cases. We found the guideline should be designed considering various factors: the infectiousness threshold values, the detection limit of antigen tests, symptom presence, and an acceptable level of releasing infectious patients. Especially, when detection limit is higher than the infectiousness threshold values, more consecutive negative results are needed to ascertain loss of infectiousness. To control the risk of releasing of infectious individuals under certain levels, rapid antigen tests should be designed to have lower detection limits than infectiousness threshold values to minimize the length of prolonged isolation, and the length of prolonged isolation increases when the detection limit is higher than the infectiousness threshold values, even though the guidelines are optimized for given conditions.

## Introduction

Vaccination campaign for COVID-19 are being successfully implemented over the world (World Health Organization). However, despite the high vaccination coverages achieved in many Western countries (World Health Organization), the emergence of the Omicron variant reminded us how vaccination alone may not be sufficient to prevent new major waves of infection (World Health Organization). Nonpharmaceutical interventions (NPIs), such as wearing masks, social distancing, reactive closures, still play a central role in the pandemic response and testing, isolation, and quarantine represent its backbone (Aleta et al., 2020).

One of the point of discussion regarding the isolation of SARS-CoV-2 infected individuals is when to end the isolation period. A longer isolation decreases the risk of transmission after the isolation; however, it may impose unnecessarily lengthy isolation, which is a burden on physical and mental health of the patients (Mian, Al-Asad, & Khan, 2021) and economy (Ash, Bento, Kaffine, Rao, & Bento, 2021). The criteria for ending isolation need to be determined considering the balance between pros and cons of the isolation.

There are two main approaches widely adapted by countries to determine the end of the isolation of COVID-19 patients. One is to isolate infected patients over a fixed time, whereas the other is to isolate infected patients until their viral load drops below a “safe(r)” level (Centers for Disease Control and Prevention, 2020). In our previous study, we demonstrated that the latter approach, based on PCR testing of isolated individuals, could minimize unnecessary isolation while controlling the risk of further transmission (Jeong et al., 2021). This is because some patients are no longer infectious by the end of isolation (thus they are redundantly isolated), whereas others may still be infectious, due to substantial individual variability in viral dynamics (Iwanami et al., 2021). However, PCR tests have a few limitations when used to determine the end of isolation. First, the turnaround time is a day or two (Larremore et al., 2021), suggesting patients need to wait a day or two until they are released from isolation even though they were not infectious anymore. Second, PCR tests are pricy. The cost of single PCR test is 51 USD (Baggett et al., 2020), whereas that of rapid antigen tests is 5 USD (Du et al., 2021) in the US, although the cost could differ between countries. Further, the facilities for PCR tests are not available everywhere.

In the US, the Centers for Disease Control and Prevention (CDC) created guidelines for when to discontinue precautions (thus isolation) for COVID-19 patients in health care settings (Centers for Disease Control and Prevention, 2020). In the early phase of the pandemic, the guideline included the use of PCR tests as follows: “Results are negative from at least two consecutive respiratory specimens collected ≥ 24 hours apart” (a test-based guideline)(Centers for Disease Control and Prevention, 2020). However, on August 10, 2020, possibly due to the discussed limitations of PCR testing, the guideline was updated as follows: “At least 10 days have passed since symptoms first appeared”, because “in the majority of cases, it [a test-based guideline] results in prolonged isolation of patients who continue to shed detectable SARS-CoV-2 RNA but are no longer infectious.” (Centers for Disease Control and Prevention, 2020).

Given these limitations of PCR tests, the use of antigen tests in determining the end of the isolation period could be considered. On one hand, antigen tests have a few advantages compared to PCR tests: i) a shorter turnaround time (less than an hour)(Butler et al., 2021; Dao Thi et al., 2020; Larremore et al., 2021; Yang et al., 2021); ii) low cost, and iii) easier accessibility. On the other hand, the low sensitivity of rapid antigen tests could be an issue. The detection limit of antigen tests is about 10^5.0^ copies/mL (Butler et al., 2021; Dao Thi et al., 2020; Miyakawa et al., 2021; Yang et al., 2021), whereas that of PCR tests is about 10^2.0^ copies/mL (Fung et al., 2020; Giri et al., 2021; van Kasteren et al., 2020). However, the infectiousness threshold values assessed by epidemiological data and in-vivo experiments (i.e., culturability) was estimated to be 10^5.0∼6.0^ (van Kampen et al., 2021; Wölfel et al., 2020), which is close to or slightly higher than the detection limits of antigen tests. This supported the use of antigen test screening to mitigate transmission (Larremore et al., 2021; Liu et al., 2022; Quilty et al., 2021).

Here, we conduct a modeling study evaluate the use of antigen tests to determine the end of the isolation period, minimizing both the risk of onward transmission following isolation and the burden of the isolation.

## Materials and Methods

### Viral load data

Longitudinal viral load data of symptomatic and asymptomatic COVID-19 patients were extracted from literatures using PubMed and Google Scholar. To estimate parameters of the viral dynamics model, we used the data satisfying the following criteria: 1) viral load was measured at three different time points at least; 2) viral load was measured from upper respiratory specimens (i.e., nose or pharynx); 3) patients were not treated with antiviral drugs or vaccinated before infection (because the model does not account for vaccine and antiviral effect). All data were collected from 2020 to early 2021, and are alpha, epsilon, and non-variants of interest/variants of concern (VOI/VOCs) as well as the original variant. As all the data used in this study were from published data and deidentified, ethics approval was not needed.

### Modeling SARS-CoV-2 viral dynamics and parameter estimation

The viral load data were used to parameterize the mathematical models of viral dynamics. The detail of the models is available in our previous study (Jeong et al., 2021). Under reasonable parameter settings, the trajectory of viral load *V*(*t*) shapes a bell-shaped curve; the viral load increases exponentially first, hit the peak, and then declines because of limited uninfected target cells (those monotonically decrease as virus increases). A nonlinear mixed-effect model was used for parameter estimation as in the previous study (Jeong et al., 2021). Model parameters were estimated independently for symptomatic and asymptomatic patients.

### Simulation of viral dynamics and ending isolation following different guidelines

True viral load data, *V*(*t*), for 1,000 patients were simulated by running the developed viral dynamics model. Parameter values of the simulation for each patient were resampled from the posterior distributions estimated in the fitting process. Accounting for measurement error (mainly due to sampling process), the measured viral load is assumed as a sum of true viral load and measurement error: 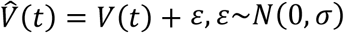, where *ε* is the measurement error term. The variance of the error term, *σ*^2^, was estimated in the fitting process. We assumed that the isolation and the first test was performed 8 days after infection. This assumption does not influence our results as this study focuses on the late phase of the infection (i.e., when the viral load reaches the detection limit of antigen tests). The test is repeated with a fixed time interval until a fixed number of consecutive negative results (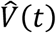< detection limit) are observed. To simulate different guidelines, we varied the time interval of tests and the number of consecutive negative results. The detection limits of the antigen test were varied from 10^4^ copies/mL to 10^6^ copies/mL. The lowest value (10^4^ copies/mL) corresponds to the antigen test kits developed by Fujifilm, and the highest value (10^6^ copies/mL) corresponds the one broadly used and developed by Abbot (Miyakawa et al., 2021). The threshold level for infectiousness is still uncertain and thus we investigated different values from 10^4.5^ copies/mL to 10^5.5^ copies/mL (Jeong et al., 2021). Simulations were separately performed for symptoamtic patients and asymptomatic patients.

### Designing the isolation guideline utilizing antigen tests

In exploring different isolation guidelines, two metrics are considered: 1. the probability of prematurely ending isolation, and 2. the length of unnecessarily prolonged isolation, both of which are defined in the previous paper (Jeong et al., 2021). For simplicity, we define the first metric as “risk”, and second metric as “burden” of isolation.

Balancing those two metrics are challenging because stricter guidelines (i.e., more consecutive negative results and longer interval of tests) contributes to reducing the risk, however, yields to unnecessarily long isolation. Therefore, the best guideline is defined as the combination of time interval of tests and consecutive negative results which controls the risk of ending isolation of infectious patients under a certain level (1% or 5%) while minimizing the prolonged isolation.

## Results

### Descriptive statistics

In total, 10 papers included at least one patient meeting the inclusion criteria. In those papers, 109 and 101 were symptomatic and asymptomatic cases, respectively. There were 85, 117, and 8 patients from Asia, USA, and Europe, respectively (**Table 1**). In most studies, cycle thresholds were reported instead of viral load. Therefore, the cycle threshold was converted to viral load (copies/mL) using the conversion formula: log_10_(Viral laod [copies/mL]) = −0.32 × Ct values [cycles] + 14.11 (Peiris et al., 2003). All the patients in those studies were hospitalized regardless of the symptom status; however, clinical course of infection (i.e., severity) was not consistently available.

**Table 1.**
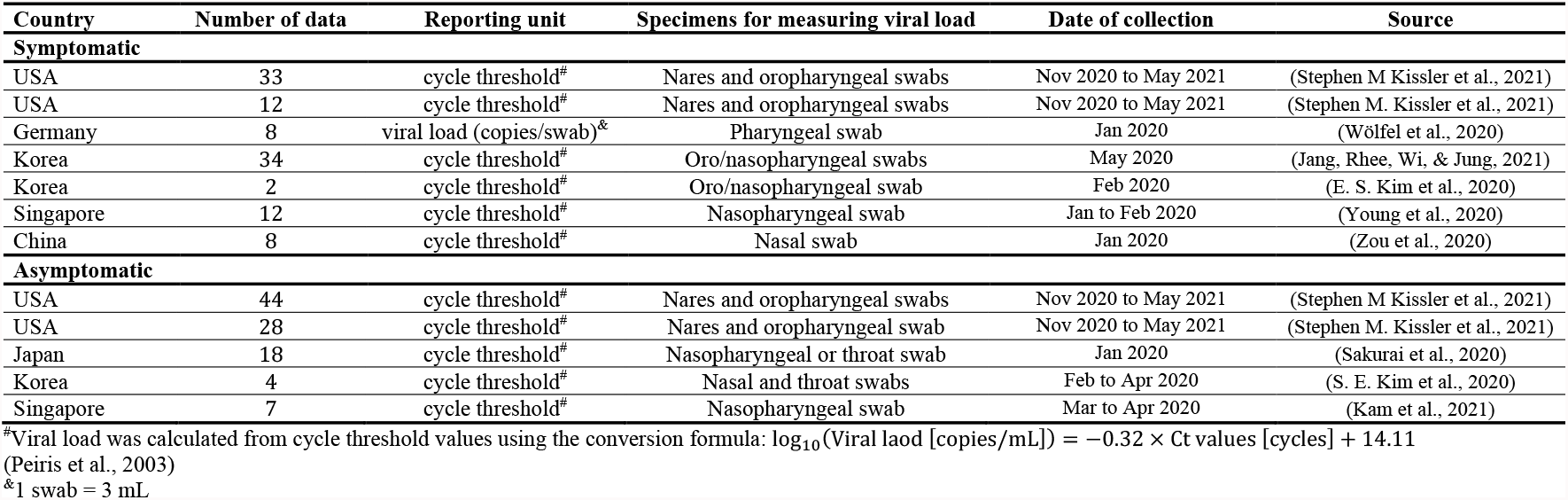
Summary of the viral load data used for modeling.

### Model fitting to the asymptomatic and symptomatic individuals

**Figure 1** shows the fitted curves of viral load for symptomatic and asymptomatic patients using estimated fixed effect parameters. For both cases, the peak viral load appears about 4 days after infection. However, the peak viral load was higher in symptomatic cases (about 10^6.5^ copies/mL for symptomatic cases vs. 10^6.0^ copies/mL for asymptomatic cases), and the viral load remained relatively high for longer time in symptomatic individuals. The viral load drops below 1 copy/mL at day 25 (95%CI: 21-29) and 21 (95%CI: 17-24) for symptomatic and asymptomatic cases, respectively. The difference on peak value of the viral load between symptomatic and asymptomatic cases was observed, which is explained by difference on the rate constant for virus infection in the model (**Supplementary File 1**). The quicker clearance of the virus in asymptomatic individuals is explained by a stronger immune response, with a higher death rate of infected cells in the model (**Supplementary File 1**). This finding is in agreement with previous studies suggesting lower viral load and shorter persistence of viral RNA in mild than in severe cases (Sun et al., 2020; Zhang et al., 2020; Zheng et al., 2020) and a longer persistence of viral RNA in symptomatic individuals (Stephen M. Kissler et al., 2021). Given these differences in the viral dynamics, we evaluate different isolation guideline for symptomatic and asymptomatic individuals.

**Figure 1.**
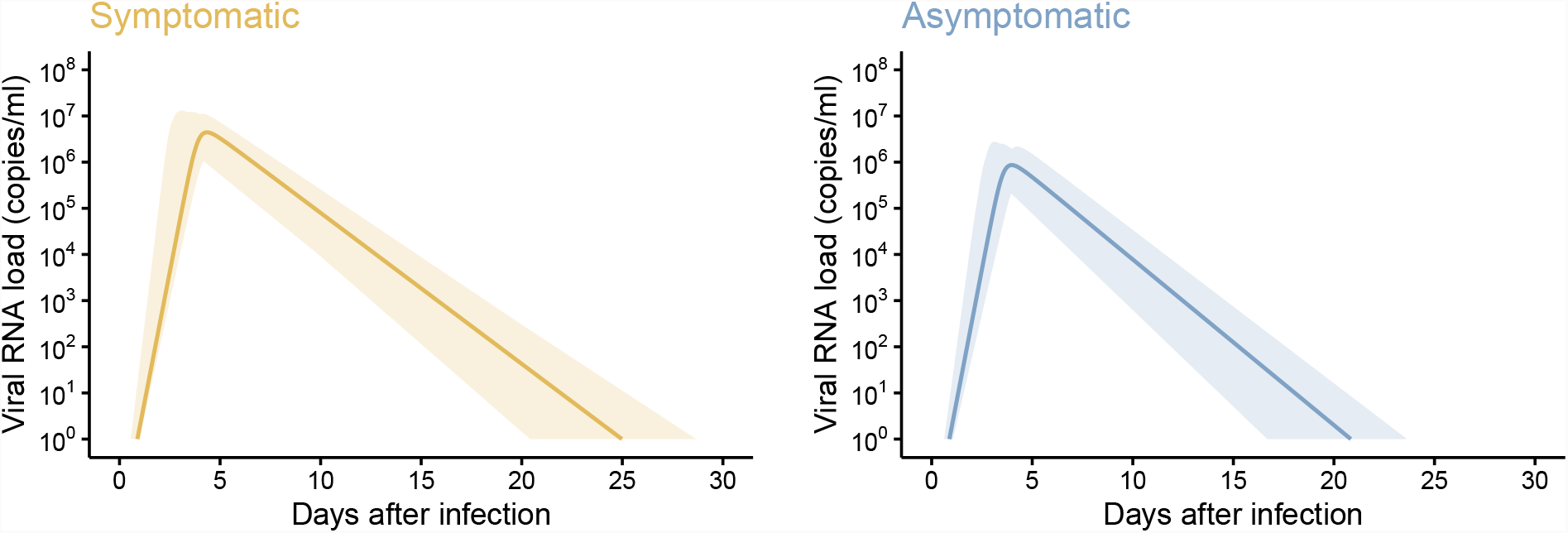
Estimated viral load curves from the models for (A) symptomatic and (B) asymptomatic cases. The solid lines are the estimated viral load curves for the best fit parameters. The shaded regions correspond to 95% predictive intervals. The 95% predictive interval was created using bootstrap approach.

### Antigen tests to end isolation

**Figures 2** and **3** show the probability of prematurely ending isolation (risk) and the length of unnecessarily prolonged isolation (burden) for symptomatic and asymptomatic cases, respectively, by varying the consecutive negative results, interval between tests, and infectiousness threshold values. The detection limit of rapid antigen tests was assumed to be 10^4^ copies/mL and 10^6^ copies/mL in **Figures 2** and **3**, respectively. As we observed in our previous paper (Jeong et al., 2021), regardless of detection limits, infectiousness threshold values, and symptom presence, the risk declined as the interval between tests becomes longer and more consecutive negative results are needed. Meanwhile, the burden increased at the same time.

**Figure 2.**
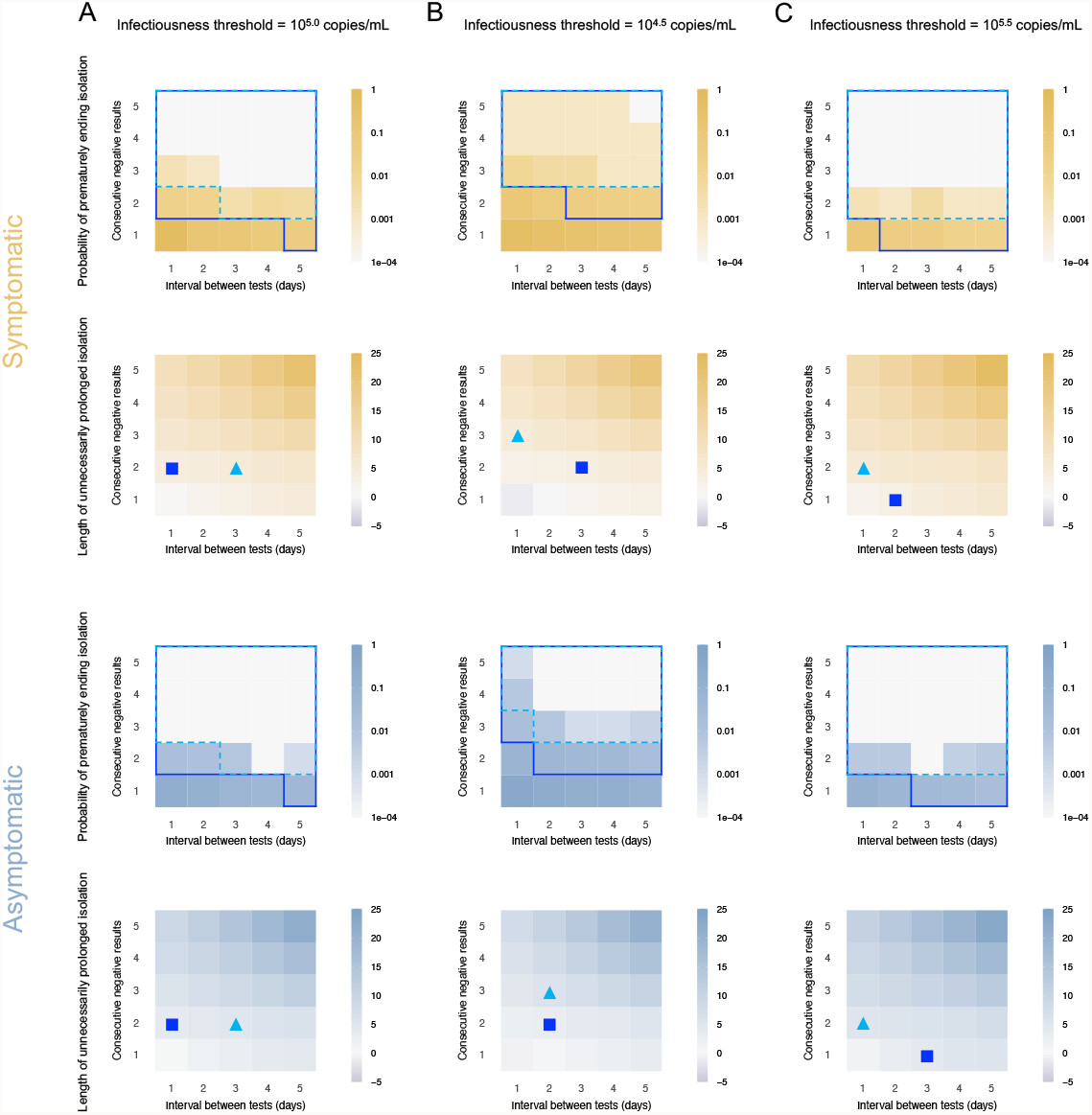
Optimal isolation guideline for symptomatic and asymptomatic cases using antigen test (detection limit=10^4^ copies/mL). **A**. Probability of prematurely ending isolation (upper panels) and mean length of unnecessarily prolonged isolation (lower panels) for different values of the interval between PCR tests and the number of consecutive negative results necessary to end isolation for each case; the infectiousness threshold value is set to 10^5.0^ copies/mL. The areas surrounded by sky-blue dotted lines and blue solid lines are those with 1% or 5% or lower of risk of prematurely ending isolation of infectious patients, respectively, and the triangles and squares correspond to the conditions which realize the shortest prolonged isolation within each area. **B**. Same as **A**, but for an infectiousness threshold value of 10^4.5^ copies/mL. **C**. Same as **A**, but for an infectiousness threshold value of 10^5.5^ copies/mL. Color keys and symbols apply to all panels.

**Figure 3.**
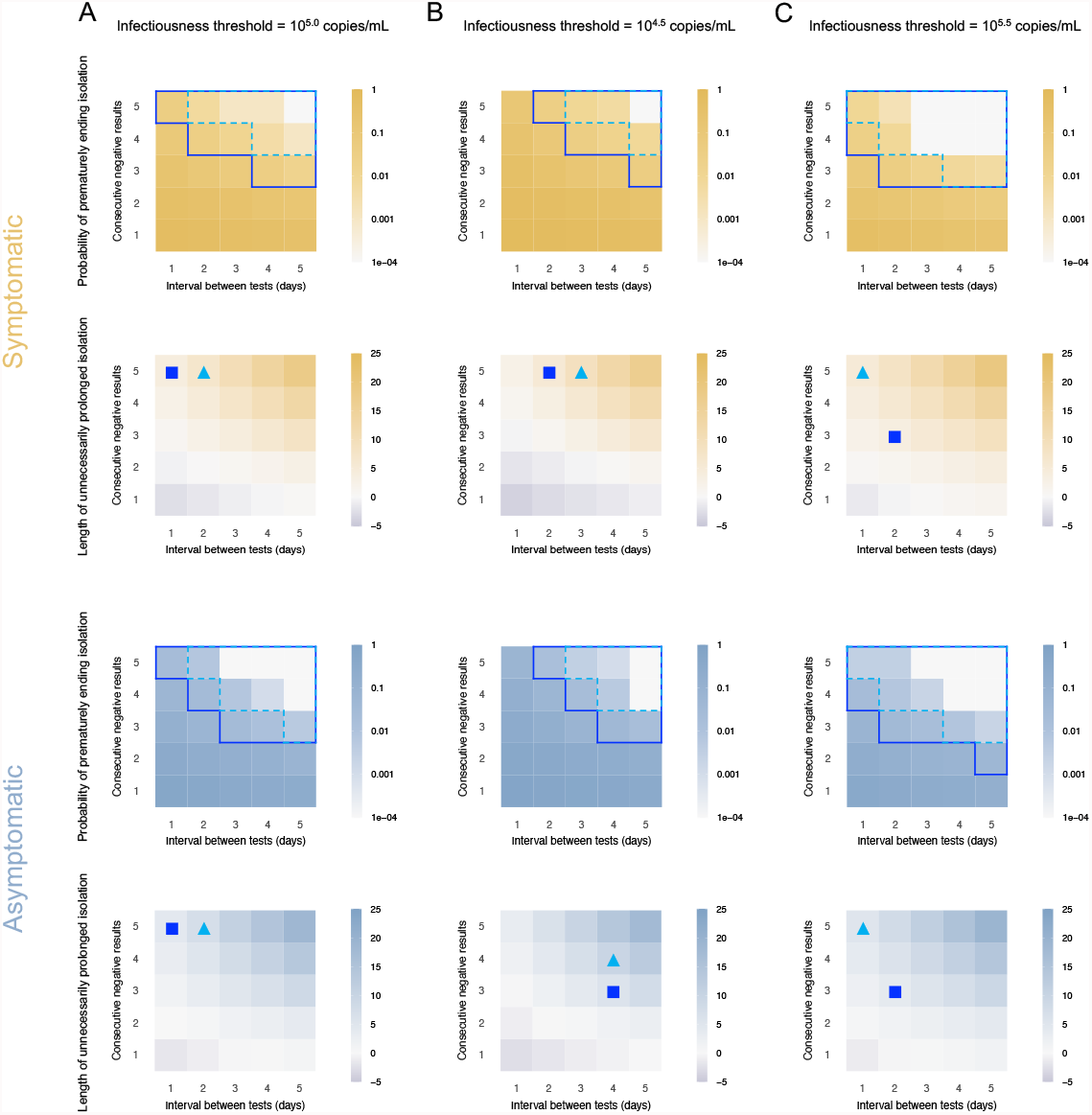
Optimal isolation guideline for symptomatic and asymptomatic cases using antigen test (detection limit=10^6^ copies/mL). **A**. Probability of prematurely ending isolation (upper panels) and mean length of unnecessarily prolonged isolation (lower panels) for different values of the interval between PCR tests and the number of consecutive negative results necessary to end isolation for each case; the infectiousness threshold value is set to 10^5.0^ copies/mL. The areas surrounded by sky-blue dotted lines and blue solid lines are those with 1% or 5% or lower of risk of prematurely ending isolation of infectious patients, respectively, and the triangles and squares correspond to the conditions which realize the shortest prolonged isolation within each area. **B**. Same as **A**, but for an infectiousness threshold value of 10^4.5^ copies/mL. **C**. Same as **A**, but for an infectiousness threshold value of 10^5.5^ copies/mL. Color keys and symbols apply to all panels.

Should 5% or lower risk of prematurely ending isolation be considered as acceptable, it is not possible to identify a single optimal strategy as the effectiveness of the guideline are estimated to depend on the infectiousness threshold, detection limits of the antigen test, and symptom presence. For example, when the detection limit and infectiousness threshold value were 10^4^ copies/mL and 10^5^ copies/mL, the optimal guideline (denoted by the square in **Figures 2** and **3**) for symptomatic individuals was to perform tests every day and to observe 2 consecutive negative results before ending the isolation (risk: 0.020 [95%CI: 0.016 to 0.025] and burden: 4.0 days [95% empirical CI: 0 to 10]). The optimal guideline also depends on the acceptable risk of prematurely ending isolation. When a 1% or lower risk is considered to be acceptable, more consecutive negative results would be needed to end isolation. When the detection limit is high (10^6^ copies/mL), an optimal guideline would require more consecutive negative results, as the infectiousness threshold values are below the detection limit and limited number of consecutive negative results cannot guarantee that the viral load is below the infectiousness threshold.

**Figure 4** summarized the burden of isolation when considering the identified optimal guideline under different conditions (i.e., symptom presence, acceptable level of risk, and infectiousness threshold values). Low burden was realized when higher risk could be accepted (comparison between **Figure 4A** and **4B**). The influence of symptom presence on the burden was estimated to be limited.

**Figure 4.**
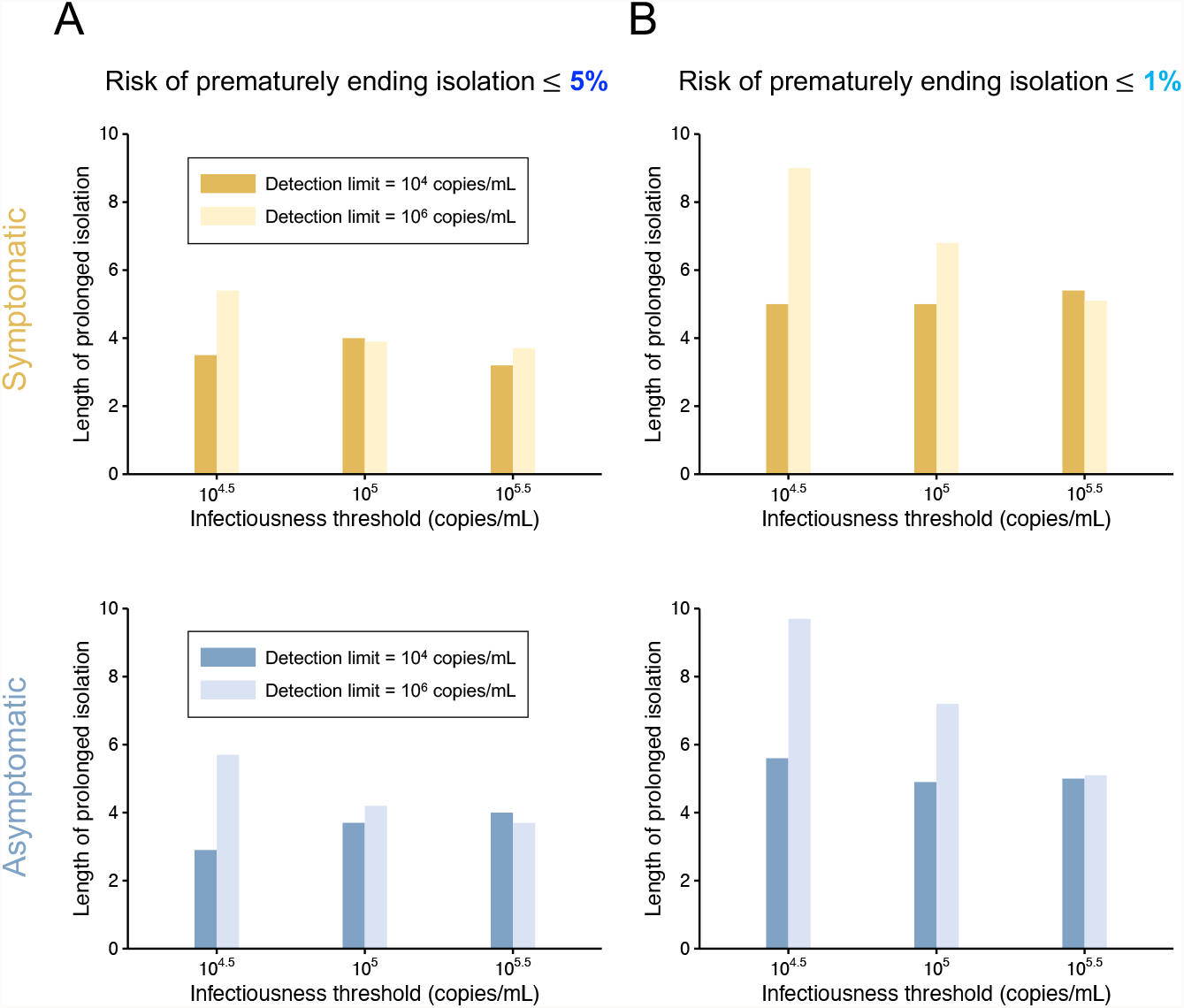
Comparison between the situations of high and low detection limits for symptomatic and asymptomatic cases. **A**. Mean length of prolonged isolation for different infectiousness threshold values and for the two approaches when considering a 5% or lower risk of prematurely ending isolation. Note that the interval between antigen tests and the number of consecutive negative results necessary to end isolation were selected to minimize the duration of prolonged isolation. **B**. Same as **A**, but considering a 1% or lower risk of prematurely ending isolation.

The influence of the combination of infectiousness threshold values and detection limits on the burden was intriguing. When the detection limit was higher than the infectiousness threshold value (i.e., detection limit was 10^6^ copies/mL), the burden was minimized when the detection limit is close to the infectiousness threshold values. However, when the detection limit is lower than the infectiousness threshold values, the burden was not much influenced by the infectiousness threshold values. That says, rapid antigen tests should have lower detection limits than infectiousness threshold values, and the burden becomes large when the detection limit is much higher than the infectiousness threshold values, even though the guidelines are optimized for given conditions.

## Discussion

We provide a quantitative assessment of alternative guidelines for the definition of duration of the isolation period based on the use of rapid antigen tests. We found that the optimal guideline was depending on the acceptable risk, detection limits, infectiousness threshold values, in agreement with what was estimated for PCR-based exit testing guidelines (Jeong et al., 2021). Among those three factors, the detection limit was positively associated with consecutive negative results necessary to end isolation. In other words, more consecutive negative results are necessary when the detection limit is above infectiousness threshold values. Our study supports the need to define different testing strategies to end the isolation for symptomatic and asymptomatic individuals. Comparing the burden of isolation (i.e., length of prolonged isolation) depending on different settings, we found rapid antigen tests should have lower detection limits than infectiousness threshold values, and the burden increases as the detection limit is much higher than the infectiousness threshold values, even though the guidelines are optimized for given conditions.

The burden of isolation under optimal guidelines was influenced by infectiousness threshold values, which was not observed in the previous study using PCR tests (Jeong et al., 2021). PCR tests can quantitatively measure viral load; thus, the measured viral load is directly compared against the infectiousness threshold value whatever the value is. Therefore, the impact of infectiousness threshold values was not observed on the burden of isolation under optimal guidelines when PCR tests are used (Jeong et al., 2021). Meanwhile, as results from rapid antigen tests are qualitative (i.e., positive, or negative), we only know whether the viral load is below the detection limit, but we do not necessarily know whether it is below the infectiousness threshold value depending on the values. For instance, if the detection limit is below the infectiousness threshold value (detection limit is 10^4^ copies/mL in this study), negative antigen tests results suggest that the viral load is below the infectiousness threshold value. In such case, we did not find much influence of infectiousness threshold values on the burden of isolation. Meanwhile, if the detection limit is above the infectiousness threshold value (detection limit is 10^6^ copies/mL in this study), negative antigen results does not necessarily suggest that the viral load is below infectiousness threshold value. Therefore, in such cases, the burden increases when the difference between the infectiousness threshold value and the detection limit is large.

A limitation of this study is that the data used to calibrate the model refers to the original SARS-CoV-22 lineage. Previous studies suggest the viral dynamics are different between the original and the Delta variant (Li et al., 2021). Moreover, we do not have data to calibrate the model for vaccinated individuals, let alone with different vaccine types and number of doses, and previous studies have shown differences in the viral load of infected vaccinated vs. infected unvaccinated individuals (Chia et al.).

The COVID-19 pandemic is having an unprecedented impact on the lives of nearly every human being on the planet and is still causing interruptions in educational and economic activities. Isolating infected individuals is still a key component of the pandemic response and development of appropriate isolation guidelines is needed. Our study provides insights on the use of rapid antigen tests to minimize both the burden of isolation and the risk of releasing infectious individuals, and suggest that different guidelines may be warranted for symptomatic and asymptomatic individuals.

## Data Availability

All data produced in the present study are available upon reasonable request to the authors.

## Acknowledgments

This study was supported in part by The Tokyo Foundation for Policy Research (to K.E. and K.S.); Grants-in-Aid for JSPS Scientific Research (KAKENHI) Scientific Research B 18KT0018 (to S.IWAMI), 18H01139 (to S.IWAMI), 16H04845 (to S.IWAMI), Scientific Research in Innovative Areas 20H05042 (to S.IWAMI), 19H04839 (to S.IWAMI), 18H05103 (to S.IWAMI); AMED CREST 19gm1310002 (to S.IWAMI); AMED Japan Program for Infectious Diseases Research and Infrastructure, 20wm0325007h0001, 20wm0325004s0201, 20wm0325012s0301, 20wm0325015s0301 (to S.IWAMI); AMED Research Program on HIV/AIDS 19fk0410023s0101 (to S.IWAMI); AMED Research Program on Emerging and Re-emerging Infectious Diseases 19fk0108156h0001, 20fk0108140s0801 and 20fk0108413s0301 (to S.IWAMI); AMED Program for Basic and Clinical Research on Hepatitis 19fk0210036h0502 (to S.IWAMI); AMED Program on the Innovative Development and the Application of New Drugs for Hepatitis B 19fk0310114h0103 (to S.IWAMI); Moonshot R&D Grant Number JPMJMS2021 (to S.IWAMI) and JPMJMS2025 (to S.IWAMI); JST MIRAI (to S.IWAMI); Mitsui Life Social Welfare Foundation (to S.IWAMI); Shin-Nihon of Advanced Medical Research (to S.IWAMI); Suzuken Memorial Foundation (to S.IWAMI); Life Science Foundation of Japan (to S.IWAMI); SECOM Science and Technology Foundation (to S.IWAMI); The Japan Prize Foundation (to S.IWAMI); Foundation for the Fusion of Science and Technology (to S.IWAMI); the MIDAS Coordination Center (MIDASSUGP2020-6) by a grant from the National Institute of General Medical Science (3U24GM132013-02S2) (to K.E. and M.A.). The study does not necessarily represent the views of the funding agencies listed above.

## Legend for figures and supplementary files

**Supplementary File 1**. Estimated parameters of SARS-CoV-2 viral dynamics model for symptomatic and asymptomatic cases.

**Figure 1-source data 1. Estimated viral load curves**. The numbers in parentheses are the 95% empirical CI.

**Figure 2-source data 1. Probability of prematurely ending isolation of infectious patients with different guidelines for symptomatic cases (with 10^5.0^ copies/mL as an infectiousness threshold value and detection limit as 10^4^ copies/mL)**. The cell with numbers in bold corresponds to the baseline. The numbers in parentheses are the 95%CI.

**Figure 2-source data 2. Length of unnecessarily prolonged isolation with different guidelines for symptomatic cases (with 10^5.0^ copies/mL as an infectiousness threshold value and detection limit as 10^4^ copies/mL)**. The cell with numbers in bold corresponds to the baseline. The numbers in parentheses are the empirical 95%CI.

**Figure 2-source data 3. Probability of prematurely ending isolation of infectious patients with different guidelines for symptomatic cases (with 10^4.5^ copies/mL as an infectiousness threshold value and detection limit as 10^4^ copies/mL)**. The cell with numbers in bold corresponds to the baseline. The numbers in parentheses are the 95%CI.

**Figure 2-source data 4. Length of unnecessarily prolonged isolation with different guidelines for symptomatic cases (with 10^4.5^ copies/mL as an infectiousness threshold value and detection limit as 10^4^ copies/mL)**. The cell with numbers in bold corresponds to the baseline. The numbers in parentheses are the empirical 95%CI.

**Figure 2-source data 5. Probability of prematurely ending isolation of infectious patients with different guidelines for symptomatic cases (with 10^5.5^ copies/mL as an infectiousness threshold value and detection limit as 10^4^ copies/mL)**. The cell with numbers in bold corresponds to the baseline. The numbers in parentheses are the 95%CI.

**Figure 2-source data 6. Length of unnecessarily prolonged isolation with different guidelines for symptomatic cases (with 10^5.5^ copies/mL as an infectiousness threshold value and detection limit as 10^4^ copies/mL)**. The cell with numbers in bold corresponds to the baseline. The numbers in parentheses are the empirical 95%CI.

**Figure 2-source data 7. Probability of prematurely ending isolation of infectious patients with different guidelines for asymptomatic cases (with 10^5.0^ copies/mL as an infectiousness threshold value and detection limit as 10^4^ copies/mL)**. The cell with numbers in bold corresponds to the baseline. The numbers in parentheses are the 95%CI.

**Figure 2-source data 8. Length of unnecessarily prolonged isolation with different guidelines for asymptomatic cases (with 10^5.0^ copies/mL as an infectiousness threshold value and detection limit as 10^4^ copies/mL)**. The cell with numbers in bold corresponds to the baseline. The numbers in parentheses are the empirical 95%CI.

**Figure 2-source data 9. Probability of prematurely ending isolation of infectious patients with different guidelines for asymptomatic cases (with 10^4.5^ copies/mL as an infectiousness threshold value and detection limit as 10^4^ copies/mL)**. The cell with numbers in bold corresponds to the baseline. The numbers in parentheses are the 95%CI.

**Figure 2-source data 10. Length of unnecessarily prolonged isolation with different guidelines for asymptomatic cases (with 10^4.5^ copies/mL as an infectiousness threshold value and detection limit as 10^4^ copies/mL)**. The cell with numbers in bold corresponds to the baseline. The numbers in parentheses are the empirical 95%CI.

**Figure 2-source data 11. Probability of prematurely ending isolation of infectious patients with different guidelines for asymptomatic cases (with 10^5.5^ copies/mL as an infectiousness threshold value and detection limit as 10^4^ copies/mL)**. The cell with numbers in bold corresponds to the baseline. The numbers in parentheses are the 95%CI.

**Figure 2-source data 12. Length of unnecessarily prolonged isolation with different guidelines for asymptomatic cases (with 10^5.5^ copies/mL as an infectiousness threshold value and detection limit as 10^4^ copies/mL)**. The cell with numbers in bold corresponds to the baseline. The numbers in parentheses are the empirical 95%CI.

**Figure 3-source data 1. Probability of prematurely ending isolation of infectious patients with different guidelines for symptomatic cases (with 10^5.0^ copies/mL as an infectiousness threshold value and detection limit as 10^6^ copies/mL)**. The cell with numbers in bold corresponds to the baseline. The numbers in parentheses are the 95%CI.

**Figure 3-source data 2. Length of unnecessarily prolonged isolation with different guidelines for symptomatic cases (with 10^5.0^ copies/mL as an infectiousness threshold value and detection limit as 10^6^ copies/mL)**. The cell with numbers in bold corresponds to the baseline. The numbers in parentheses are the empirical 95%CI.

**Figure 3-source data 3. Probability of prematurely ending isolation of infectious patients with different guidelines for symptomatic cases (with 10^4.5^ copies/mL as an infectiousness threshold value and detection limit as 10^6^ copies/mL)**. The cell with numbers in bold corresponds to the baseline. The numbers in parentheses are the 95%CI.

**Figure 3-source data 4. Length of unnecessarily prolonged isolation with different guidelines for symptomatic cases (with 10^4.5^ copies/mL as an infectiousness threshold value and detection limit as 10^6^ copies/mL)**. The cell with numbers in bold corresponds to the baseline. The numbers in parentheses are the empirical 95%CI.

**Figure 3-source data 5. Probability of prematurely ending isolation of infectious patients with different guidelines for symptomatic cases (with 10^5.5^ copies/mL as an infectiousness threshold value and detection limit as 10^6^ copies/mL)**. The cell with numbers in bold corresponds to the baseline. The numbers in parentheses are the 95%CI.

**Figure 3-source data 6. Length of unnecessarily prolonged isolation with different guidelines for symptomatic cases (with 10^5.5^ copies/mL as an infectiousness threshold value and detection limit as 10^6^ copies/mL)**. The cell with numbers in bold corresponds to the baseline. The numbers in parentheses are the empirical 95%CI.

**Figure 3-source data 7. Probability of prematurely ending isolation of infectious patients with different guidelines for asymptomatic cases (with 10^5.0^ copies/mL as an infectiousness threshold value and detection limit as 10^6^ copies/mL)**. The cell with numbers in bold corresponds to the baseline. The numbers in parentheses are the 95%CI.

**Figure 3-source data 8. Length of unnecessarily prolonged isolation with different guidelines for asymptomatic cases (with 10^5.0^ copies/mL as an infectiousness threshold value and detection limit as 10^6^ copies/mL)**. The cell with numbers in bold corresponds to the baseline. The numbers in parentheses are the empirical 95%CI.

**Figure 3-source data 9. Probability of prematurely ending isolation of infectious patients with different guidelines for asymptomatic cases (with 10^4.5^ copies/mL as an infectiousness threshold value and detection limit as 10^6^ copies/mL)**. The cell with numbers in bold corresponds to the baseline. The numbers in parentheses are the 95%CI.

**Figure 3-source data 10. Length of unnecessarily prolonged isolation with different guidelines for asymptomatic cases (with 10^4.5^ copies/mL as an infectiousness threshold value and detection limit as 10^6^ copies/mL)**. The cell with numbers in bold corresponds to the baseline. The numbers in parentheses are the empirical 95%CI.

**Figure 3-source data 11. Probability of prematurely ending isolation of infectious patients with different guidelines for asymptomatic cases (with 10^5.5^ copies/mL as an infectiousness threshold value and detection limit as 10^6^ copies/mL)**. The cell with numbers in bold corresponds to the baseline. The numbers in parentheses are the 95%CI.

**Figure 3-source data 12. Length of unnecessarily prolonged isolation with different guidelines for asymptomatic cases (with 10^5.5^ copies/mL as an infectiousness threshold value and detection limit as 10^6^ copies/mL)**. The cell with numbers in bold corresponds to the baseline. The numbers in parentheses are the empirical 95%CI.

**Figure 4-source data. Mean length of unnecessarily prolonged isolation (days) with different guidelines and infectiousness values controlling the risk of prematurely ending isolation** ≤ **5% and** ≤ **1% for symptomatic and asymptomatic cases**.

