## Supplementary File 1 for "Designing isolation guidelines for COVID-19 patients utilizing rapid antigen tests: a simulation study using viral dynamics models"

**Supplementary File 1. Estimated parameters of SARS-CoV-2 viral dynamics model**

| **Parameters** | **Symbol** | **Unit** | **Symptomatic** | **Asymptomatic** |
| --- | --- | --- | --- | --- |
| Maximum rate constant for viral replication | $\gamma$ | day^-1^ | $6.81$ | $6.06$ |
| Rate constant for virus infection | $\beta$ | (copies/mL)^-1^ day^-1^ | $9.41\times{10}^{-7}$ | $4.15\times{10}^{-6}$ |
| Death rate of infected cells | $\delta$ | day^-1^ | $0.74$ | $0.83$ |
